# Systemic hypoferraemia and severity of hypoxaemic respiratory failure in COVID-19

**DOI:** 10.1101/2020.05.11.20092114

**Authors:** Akshay Shah, Joe N. Frost, Louise Aaron, Killian Donovan, Stuart R. McKechnie, Simon J. Stanworth, Hal Drakesmith

**Author notes:** **Correspondence to:** Dr Akshay Shah, **Address:** Radcliffe Department of Medicine, Level 4 Academic Block, University of Oxford, John Radcliffe Hospital, Headley Way, Oxford, OX3 9DU, United Kingdom., **Email:**.

## Abstract

Coronavirus disease 2019 (COVID-19) mortality is associated with hypoxaemia, multiorgan failure, and thromboinflammation. However severity of disease varies considerably and understanding physiological changes that may link to poor outcomes is important. Although increased serum ferritin has been observed in COVID-19 patients consistent with inflammation, other iron parameters have not been examined to our knowledge. Because iron is required for immunity and oxygen utilisation, and dysregulated iron homeostasis has been observed in COPD, we investigated serum iron concentrations in 30 patients with COVID-19 requiring ICU admission. All patients had low serum iron but patients with severe hypoxemic respiratory failure had more profound hypoferraemia. The area under the curve for receiver operating characteristic curves for serum iron to identify severe hypoxemia was 0·95; the optimal Youden Index for distinguishing between severe and non-severe hypoxemia was a serum iron concentration of 2·9 μmol/L. By linear regression, serum iron was associated with lymphocyte count and PaO_2_/FiO_2_. In conclusion, profound hypoferraemia identifies COVID-19 patients with severe hypoxaemia. Serum iron is a simple biomarker that could be usefully employed to stratify patients and monitor disease. Severe hypoferraemia may plausibly impair critical iron-dependent processes such as lymphocyte responses and hypoxia sensing, contributing to pathology, and is potentially treatable.

---

COVID-19 caused by severe acute respiratory coronavirus 2 (SARS-CoV-2) was declared a pandemic on March 11, 2020, with the major causes of mortality being progressive hypoxia or multiorgan dysfunction^1^. Previous work has identified risk factors associated with respiratory failure in patients with COVID-19 which include older age, neutrophilia, elevated inflammatory and coagulation markers, and hyperferritinaemia^2^. Inflammation in general is often accompanied by systemic hypoferraemia^3^, and systemic hypoferraemia disturbs normal human responses to hypoxia-induced induced lung injury^4^, which is of considerable concern in patients with COVID-19. However, although ferritin concentrations in COVID-19 are well reported, there are no published data on other markers of iron status. Therefore, we sought to characterise iron parameters, including serum iron, in COVID-19 intensive care unit (ICU) patients and relate these to disease severity.

We retrospectively evaluated any serum iron profiles that were measured in critically ill patients with COVID-19 within 24 hours of admission to the ICU, John Radcliffe Hospital, Oxford, UK between March 31, 2020 and April 25, 2020. Relevant clinical and laboratory data were extracted from routinely collected healthcare records. We stratified patients according to severity of hypoxaemic respiratory failure on admission to ICU - severe (PaO_2_/FiO_2_ ratio < 100 mmHg) versus non-severe (PaO_2_/FiO_2_ ratio 100-300 mmHg)^5^. Research ethics committee approval was not required for this study as per the UK Health Research Authority Decision tool (http://www.hra-decisiontools.org.uk/research/); the local institutional board (Oxford University Hospitals NHS Foundation Trust Research and Development) approved ethical oversight and waiver of consent. No direct patient identifiable data were collected.

A total of 30 patients were included and their demographic, clinical and laboratory characteristics are shown in **Supplementary Table 1**. Overall, 17 (57%) patients were male. The median age was 57 years (IQR, 52-64). Compared with patients with non-severe hypoxaemia, patients with severe hypoxaemia had significantly lower levels of serum iron (median 2·3 (IQR, 2·2-2·5) vs 4·3 (IQR, 3·3-5·2) μmol/L, p < 0·001) and lymphocyte counts (mean (SD) 0·50 (0·2) vs. 0·87 (0·4), p = 0·015). There were no statistically significant differences in transferrin saturation and serum ferritin levels between groups **(Figure 1A)**. The area under the curve for receiver operating characteristic curves for serum iron to identify severe hypoxaemia was 0·95; the optimal Youden Index for distinguishing between severe and non-severe hypoxaemia was a serum iron concentration of 2·9 μmol/L **(Figure 1B)**. By linear regression, serum iron was associated with lymphocyte count and PaO_2_/FiO_2_ **(Figures 1C and 1D)**.

**Figure 1:**
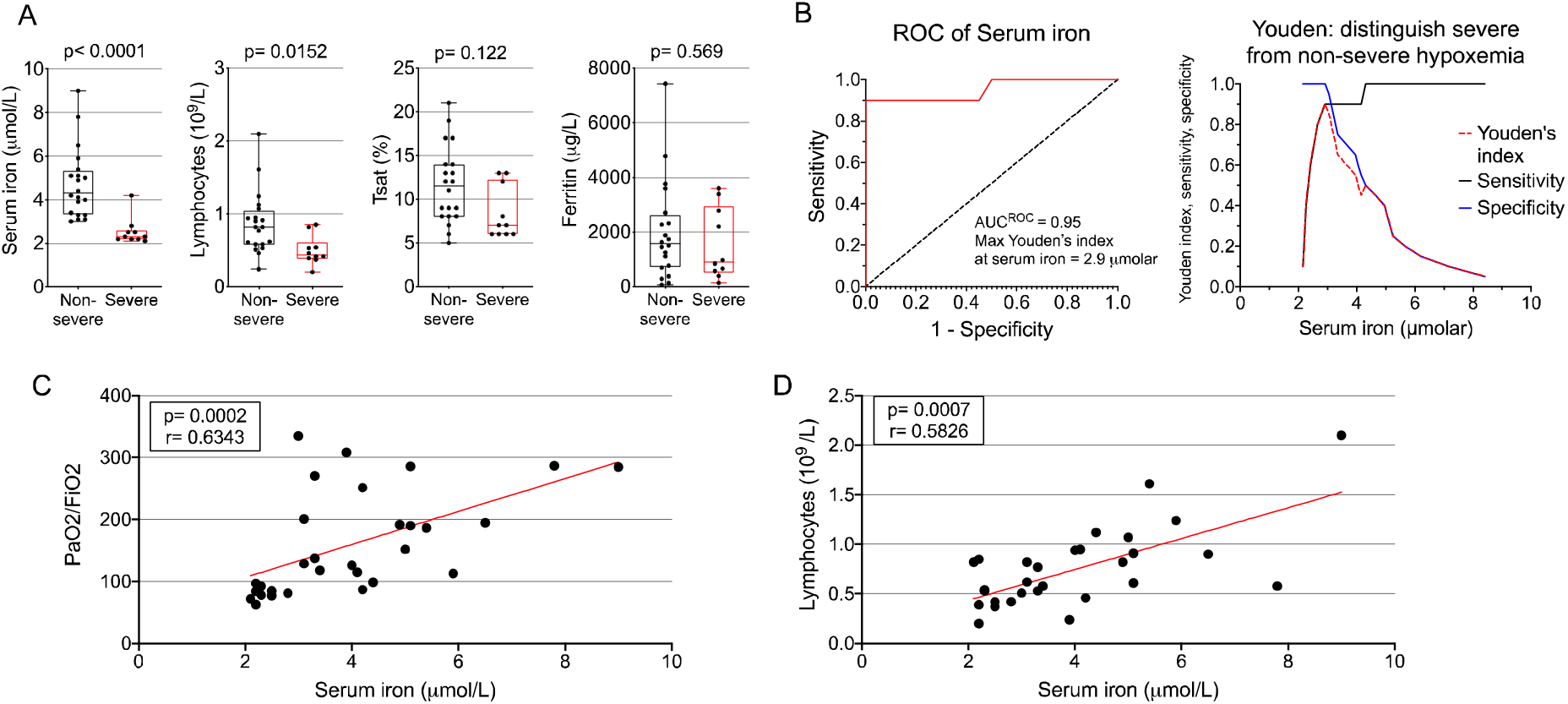
Associations between markers of iron status, lymphocyte count and severity of hypoxaemia. **Figure legend: 1A** Boxplotsshow the 25th, 50th, and 75th percentiles (box); 10th and 90th percentiles (whiskers); and data points (circles) of serum iron, transferrin saturation (Tsat) and serum ferritin, stratified by severity of hypoxemia; **IB** Receiver Operating Characteristic (ROC) Curve and Youden Index for serum iron in distinguishing severe and non-severe hypoxemia; **1C** Correlation serum iron and PaO_2_/FiO_2_ ratio; **ID** Correlation between serum iron and lymphocyte count. Mann-Whitney rank sum test was used to compare nonparametric continuous variables between these two groups. All statistical tests were 2-tailed, and statistical significance was defined as *PQ*<*Q*.05. Analyses were performed using PRISM version 8 (GraphPad Software).

Our findings demonstrate profound hypoferraemia in COVID-19 patients with severe hypoxaemia. Serum iron was lower when compared with other cohorts of non-COVID-19 ICU patients reported previously, including those with sepsis ^6^. Altered iron fluxes have been linked with ICU outcomes previously, and our data indicate that severe hypoferraemia may be detrimental in COVID-19.^6, 7^ The association of serum iron with lymphocyte counts could reflect the requirement of the adaptive immune response for iron^8^ and may be relevant for immune competence in COVID-19.

The cause of hypoferremia is likely to be due to increased hepcidin secondary to inflammation^3^. Unlike hepcidin, serum iron is measured widely and so could assist with identification and monitoring of severity of disease. Anti-inflammatory drugs such as Tocilizumab may increase serum iron by suppressing hepcidin synthesis, through inhibition of interleukin-6 (which stimulates hepcidin)^9^. In summary, a pathological role of severe hypoferraemia in COVID-19 is plausible, for example via effects on hypoxia sensing and immunity, and potentially treatable, via hepcidin inhibition.

## Data Availability

Requests for data should be submitted to the corresponding author and will be honoured.

## Author contributions

Dr Shah and Prof Drakesmith had full access to all of the anonymised data in the study and take responsibility for the integrity of the data and the accuracy of the data analysis.

*Study concept and design:* A. Shah, J.N. Frost, H. Drakesmith

*Acquisition, analysis, or interpretation of data:* All authors

*Drafting of manuscript:* A. Shah, H. Drakesmith

*Critical revision of the manuscript for important intellectual content:* All authors

*Statistical analysis:* A. Shah, J.N. Frost

## Funding / support

No specific funding was required for this study. A. Shah is currently supported by an NIHR Doctoral Research Fellowship (NIHR-DRF-2017-10-094). H. Drakesmith is supported by MRC UK award no. MC_UU_12010/10 and the Oxford NIHR Biomedical Research Centre.

---

**Supplementary Table 1.** Clinical and laboratory characteristics of study cohort, Total and stratified by severity of hypoxaemia

| Characteristic | All patients (n=30) | Severe (n=10) | Non-severe (n=20) |
| --- | --- | --- | --- |
| Age, median (IQR), y | 57 (52-64) | 57 (53-75) | 57 (52-64) |
| Males, n (%) | 17 (57) | 5 (50) | 12 (60) |
| Females, n (%) | 13 (33) | 5 (50) | 8 (40) |
| Clinical Frailty Scale, n (%) |  |  |  |
| 1 | 23 (77) | 9 (90) | 14 (70) |
| 2 | 4 (13) | 1 (10) | 3 (15) |
| >3 | 3 (10) | 0 | 3 (15) |
| Respiratory support, n (%) |  |  |  |
| Non-invasive ventilation | 18 (60) | 7 (70) | 11 (55) |
| Invasive ventilation | 26 (87) | 10 (100) | 16 (80) |
| Prone position | 17 (57) | 10 (100) | 7 (35) |
| Advanced cardiovascular support, n (%) | 4 (13) | 0 (0) | 4 (20) |
| Advanced renal support, n (%) | 10 (33) | 5 (50) | 5 (25) |
| PaO<sub>2</sub> / FiO<sub>2</sub> ratio, median (IQR) | 127.5 (87-200.6) | 82.5 (77-87) | 190.8 (127.5-277.5) |
| Laboratory data (normal range) |  |  |  |
| Haemoglobin (g/L) (120 – 150), mean (SD) | 130.4 (20.1) | 124.7 (16.7) | 133.2 (21.4) |
| White cell count (x 10<sup>9</sup>/L) (4.0 – 11.0), mean (SD) | 10.6 (4.8) | 11.0 (5) | 10.4 (4.7) |
| Lymphocyte count (x 10<sup>9</sup>/L) (1.0 – 4.0), mean (SD) | 0.74 (0.4) | 0.50 (0.2) | 0.87 (0.42) |
| D-Dimer (ug/mL) (0 - 500), median (IQR) | 3286 (1302-14227) | 9505.5 (845-5023) | 2462 (1453-9850) |
| Fibrinogen (g/L) (1.5 – 4.0), median (IQR) | 6 (5.5-6.3) | 6 (5.5-6.3) | 6.1 (5.5-6.4) |
| CRP (mg/L) (0 – 5), mean (SD) | 246.2 (100.1) | 235.8 (101.8) | 251.4 (101.5) |
| Iron parameters (normal range) |  |  |  |
| Ferritin (mcg/l) (10 - 200), median (IQR) | 1476.1 (656.6-2698) | 903.8 (566.9-2789.2) | 1566.1 (729-2511.5) |
| Serum iron(umol/L) (11 - 30), median (IQR) | 3.6 (2.5-5) | 2.3 (2.2-2.5) | 4.3 (3.3-5.2) |
| Transferrin (g/L) (1.8 – 3.6), median (IQR) | 1.5 (1.1-1.8) | 1.3 (0.8-1.8) | 1.5 (1.1-1.8) |
| Transferrin saturation (%) (16 – 50), median (IQR) | 9 (7-13) | 7 (6-12) | 12 (8-14) |
| Outcome as of 25 Apr 2020, n (%) |  |  |  |
| Died in ICU | 6 (20) | 5 (50) | 1 (5) |
| Still alive in ICU | 16 (53) | 3 (30) | 13 (65) |
| Discharged alive from ICU | 8 (27) | 2 (20) | 6 (30) |
| ICU length of stay, median (IQR), days | 8 (4-11) | 7 (4-9) | 9 (4-12) |

Abbreviations: CRP, C-reactive protein; ICU, intensive care unit; IQR, interquartile range, SD, standard deviation.

## Notes

### Competing Interest Statement

HD has received research funding from Pfizer and La Jolla Pharmaceutical Company and honoraria from
Pharmacosmos, all unrelated to this work.

### Clinical Trial

not a prospective study

